# Investigating the potential effect of topiramate on anorexia nervosa and binge eating behaviour using drug-target Mendelian randomization

**DOI:** 10.64898/2026.11.05.26367397

**Authors:** Kym-Mai Nguyen, Eva Yi-Hsien Chang, Brian Kim, Paul Ebert, Jiayue-Clara Jiang, Liang-Dar Hwang

**Affiliations:** Institute for Molecular Bioscience, The University of Queensland, St Lucia, QLD, Australia; School of Environment, The University of Queensland, St Lucia, QLD, Australia; Monell Chemical Senses Center, Philadelphia, PA, USA

**Keywords:** topiramate, anorexia nervosa, binge-eating behaviour, Mendelian randomization, drug-target Mendelian randomization, epilepsy, psychiatric disorders

## Abstract

Topiramate is an antiseizure medication used to treat epilepsy and migraine and is commonly prescribed for appetite suppression and weight loss. However, whether its pharmacological actions influence eating disorder risk remains unclear. We applied drug-target Mendelian randomization (MR) to investigate potential causal effects of topiramate on the risk of anorexia nervosa (AN; 24,223 cases and 1,243,971 controls) and binge-eating behaviour (BE; 39,279 cases and 1,227,436 controls) and, exploratorily, eight other psychiatric disorders: attention-deficit/hyperactivity disorder, anxiety disorders, alcohol use disorder, bipolar disorder, major depressive disorder, panic disorder, post-traumatic stress disorder, and schizophrenia. We used summary-data-based Mendelian randomization (SMR) to investigate genetically predicted pharmacological actions of topiramate using brain- and blood-derived *cis*-expression quantitative trait loci for genes encoding its molecular targets. Five genetic instruments consistent with topiramate’s antiepileptic action were validated: *CA4* and *CACNB3* expression in the brain and *CA4, GRIK1,* and *GRIK3* expression in the blood. Robustness of the findings was assessed using SMR-multi, HEIDI, and colocalization analyses. After multiple-testing correction, genetically predicted inhibition of *CA4* expression in blood was associated with decreased BE risk (OR = 0.867, 95% CI = 0.792–0.950, p = 0.002) and remained significant in SMR-multi (p = 0.0049). Colocalization provided moderate support for a shared causal variant underlying *CA4* expression and BE (PP_H4_ = 0.58). No robust evidence was identified for AN or the eight exploratory psychiatric outcomes. Overall, our findings warrant further clinical and functional investigations into mechanisms underlying topiramate-associated appetite suppression and weight loss and their relevance to eating-disorder risk.

## Introduction

Topiramate is a commonly prescribed second-generation, centrally acting antiepileptic drug (AED) for epilepsy and migraine prevention (1). Its antiepileptic effects are thought to involve several mechanisms, including blockade of voltage-gated sodium and calcium channels, enhancement of gamma-aminobutyric acid (GABA)-mediated neurotransmission, inhibition of glutamate-mediated neurotransmission, and inhibition of carbonic anhydrases (CAs) (2).

Beyond its approved indications, topiramate has been investigated in clinical trials and used off-label, either as monotherapy or adjunctive therapy, for obesity (3), binge-eating disorder (4), bulimia nervosa (5), post-traumatic stress disorder (PTSD) (6), bipolar disorder (BD) (7), schizophrenia (SCZ) (8), and alcohol use disorder (AUD) (9). However, the mechanisms underlying these broader therapeutic effects remain incompletely understood.

Topiramate is also associated with a wide range of adverse effects, including cognitive impairment (10), paraesthesia (11,12), fatigue (13), dizziness (12), ocular complications (14), loss of appetite (15), and weight loss. Its weight-reducing effect is relatively distinct (16), with weight loss reported in 6%-16% of newly diagnosed patients with epilepsy initiated on topiramate (17). Clinical reports have also linked topiramate use to the emergence or worsening of AN-related behaviours, including restrictive eating and self-induced vomiting, as well as misuse for weight control among individuals with AN (18–20). However, evidence for a causal relationship with AN remains limited (20,21).

In contrast, topiramate has been investigated as a treatment for binge eating disorder, and has been associated with reduced binge-eating frequency and body weight (22,23), although evidence has only been studied in the context of short-term use. Thus, long-term efficacy and safety remain unclear (24,25). AN and binge-eating behaviour (BE) have contrasting clinical presentations but share substantial genetic architecture (r_g_ = 0.46), while differing in their genetic associations with anthropometric, metabolic and impulse-control traits (26). Investigating topiramate-related target modulation across AN and BE may therefore provide complementary insights into its effect on eating-related phenotypes.

Determining whether observed drug effects are causal is challenging using conventional, observational studies. Randomized controlled trials may have insufficient duration or sample size to detect uncommon adverse effects, while post-marketing surveillance is susceptible to under-reporting, confounding, and reporting biases (27–30). Moreover, trials designed specifically to investigate potentially harmful adverse effects may be impractical or unethical (31).

Mendelian randomization (MR) uses genetic variants associated with an exposure as instrumental variables to estimate the potential causal effect on an outcome (32). MR has increasingly been applied to investigate pharmacological effects using genetic variants associated with the expression of drug-target genes (expression quantitative trait loci [eQTLs]) or protein levels (protein quantitative trait loci [pQTLs]) to approximate perturbation of the corresponding drug target (33). Leveraging large publicly available genetic datasets, drug-target MR provides an efficient and cost-effective approach for drug target prioritization (34,35), validation (33,36), repositioning (37,38), and prediction of unintended adverse events (39,40).

This study applied two-sample drug-target MR to assess whether genetically predicted modulation of established topiramate targets influences the risk of AN and BE, with exploratory analyses of eight additional psychiatric outcomes: attention-deficit/hyperactivity disorder (ADHD), anxiety disorders (ANX), AUD, BD, major depressive disorder (MDD), panic disorder (PD), PTSD, and SCZ. Genetic instruments were identified from established pharmacological targets of topiramate and validated against epilepsy risk in directions consistent with topiramate’s therapeutic action.

## Methods

A summary of the study design is presented in **Figure 1**.

**Figure 1.**
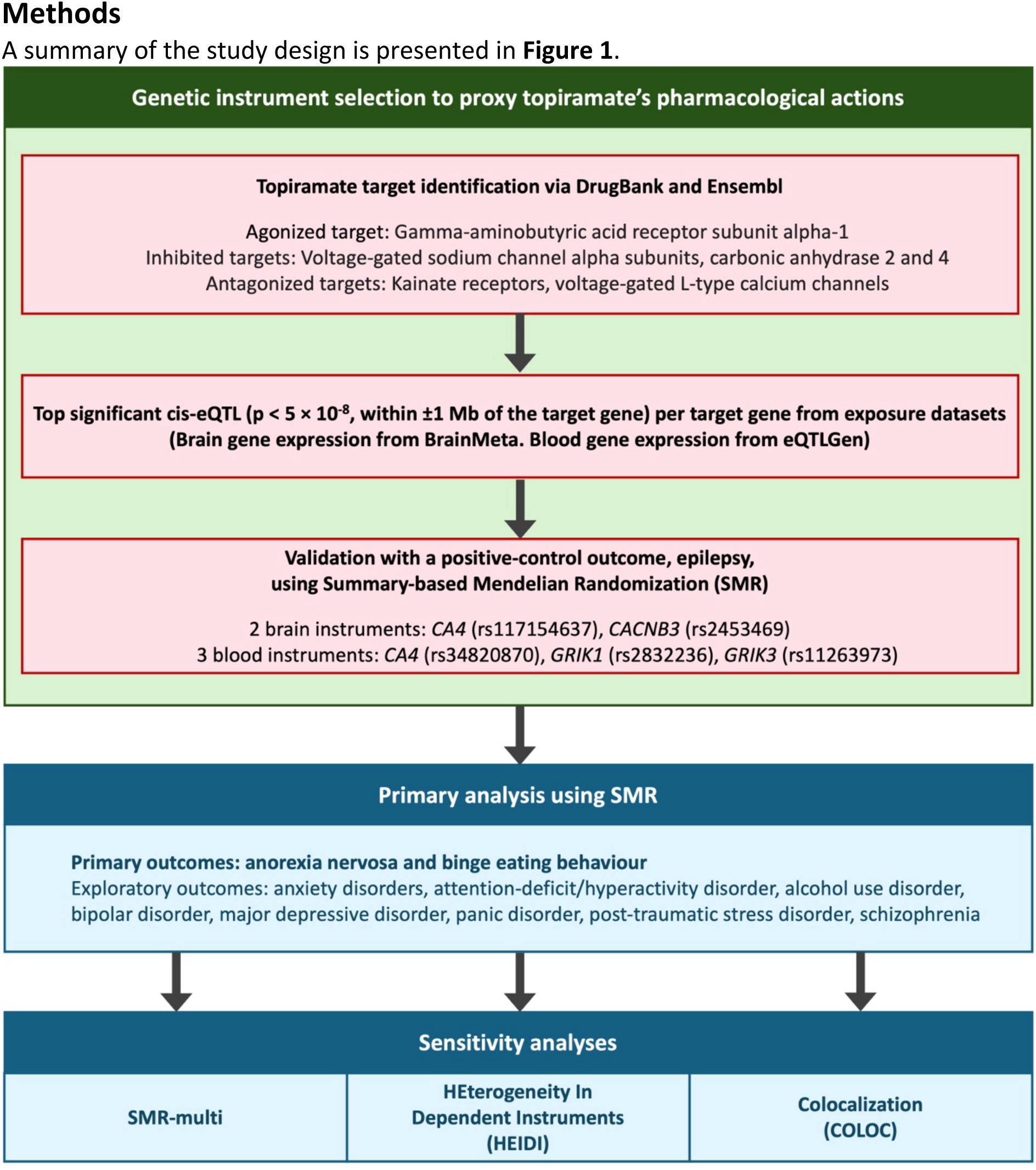
Overview of the study design.

### Data sources

Pharmacologically active molecular targets of topiramate and their corresponding encoding genes were identified through DrugBank (41) and Ensembl (42), respectively. These include gamma-aminobutyric acid receptor subunit alpha-1 (*GABRA1*), on which topiramate acts as an agonist, and 25 genes encoding targets inhibited or antagonized by topiramate: voltage-gated sodium channel alpha subunits (*SCN1A, SCN2A, SCN3A, SCN4A, SCN5A, SCN7A, SCN8A, SCN9A, SCN10A, SCN11A*), kainate receptors (*GRIK1, GRIK2, GRIK3, GRIK4, GRIK5*), carbonic anhydrase 2 (*CA2*), carbonic anhydrase 4 (*CA4*), and voltage-gated L-type calcium channels (*CACN1C, CACN1D, CACN1S, CACN1F, CACNB1, CACNB2, CACNB3, CACNB4*) (**Table 1**).

**Table 1.**
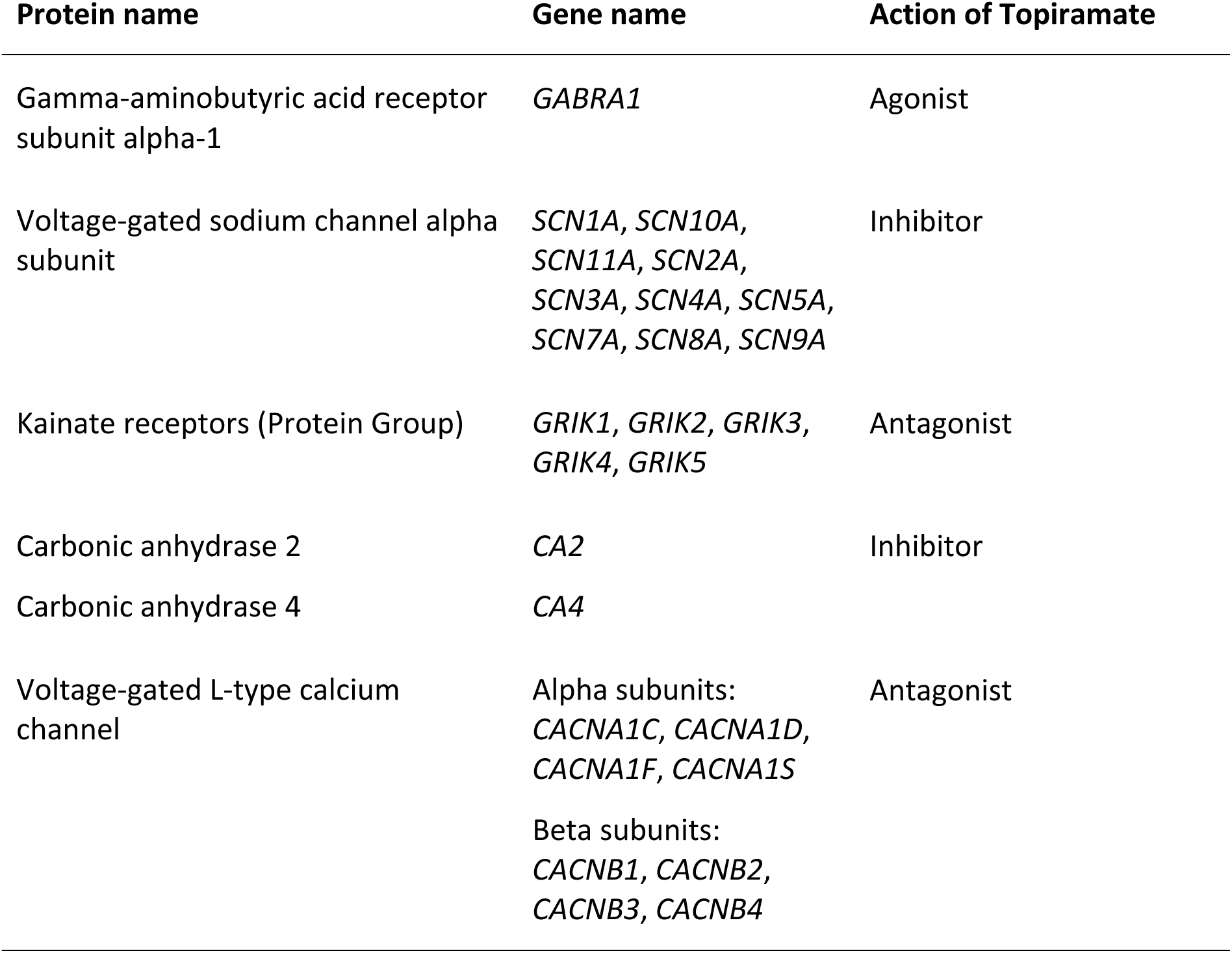
Molecular targets of topiramate, their encoding genes, and the actions of topiramate.

| Protein name | Gene name | Action of Topiramate |
|---|---|---|
| Gamma-aminobutyric acid receptor subunit alpha-1 | *GABRA1* | Agonist |
| Voltage-gated sodium channel alpha subunit | *SCN1A*, *SCN10A*, *SCN11A*, *SCN2A*, *SCN3A*, *SCN4A*, *SCN5A*, *SCN7A*, *SCN8A*, *SCN9A* | Inhibitor |
| Kainate receptors (Protein Group) | *GRIK1*, *GRIK2*, *GRIK3*, *GRIK4*, *GRIK5* | Antagonist |
| Carbonic anhydrase 2 | *CA2* | Inhibitor |
| Carbonic anhydrase 4 | *CA4* |  |
| Voltage-gated L-type calcium channel | Alpha subunits: *CACNA1C*, *CACNA1D*, *CACNA1F*, *CACNA1S*<br>Beta subunits: *CACNB1*, *CACNB2*, *CACNB3*, *CACNB4* | Antagonist |

Brain-derived *cis*-eQTL were obtained from BrainMeta (43), a genome-wide eQTL study based on RNA-seq data of 2,865 brain cortex samples from 2,443 unrelated individuals of European ancestry. Blood-derived *cis*-eQTL were obtained from eQTLGen phase I (44), comprising 31,684 samples predominantly from individuals of European ancestry. Outcomes were obtained from publicly available genome-wide association study (GWAS) summary statistics from cohorts of European ancestry, including AN (26), BE (26), and epilepsy (45). Eight additional psychiatric outcomes—ANX (46), ADHD (47), AUD (48), BD (49), MDD (50), PD (51), PTSD (52), SCZ (53)—were included in exploratory analyses. Details of the GWAS datasets are summarised in **Supplementary Table 1**.

### Instrumental variable selection

Valid genetic instrumental variables for MR should satisfy three principal assumptions: (i) the genetic variant is robustly associated with the exposure (relevance); (ii) there is no confounding association between the genetic variant and the outcome (independence); and (iii) the genetic variant influences the outcome only through the exposure and not through any pleiotropic pathway (exclusion restriction). To minimize the risk of the third MR assumption being violated, we restricted instruments to *cis*-eQTL, defined as variants located within 1 million base pairs of the corresponding gene, to reduce the potential for horizontal pleiotropy through effects on distal genes. To satisfy the relevance assumption, we selected the strongest *cis*-eQTLs associated with the expression of genes encoding molecular targets of topiramate in the brain or blood with p < 5×10^-8^. Instrument strength was assessed using the F-statistic, with an F-statistic > 10 considered indicative of sufficient instrument strength. All *cis*-eQTL and outcome datasets were restricted to individuals predominantly or entirely of European ancestry to minimize the risk of violating the second assumption through population stratification.

To ensure that the selected *cis*-eQTL reflected the pharmacological action of topiramate, we assessed epilepsy as a positive control outcome in the summary-based Mendelian randomization (SMR) analysis (54), to determine whether genetically predicted expression of each target gene was associated with epilepsy risk in the expected direction. A total of five *cis*-eQTLs showing an association with epilepsy in the expected direction (p < 0.05) were considered validated instruments and carried forward to the primary and exploratory analyses.

### Statistical analysis

We performed SMR to investigate the potential causal effect of topiramate’s genetically predicted pharmacological actions. SMR is an extension of MR that integrates summary-level data from GWAS and eQTL studies to test whether genetically predicted gene expression is associated with an outcome (54,55). Briefly, SMR uses the Wald ratio estimator (*̂β_MR_*) to estimate the causal effect 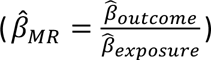 of the genetically predicted gene expression on the outcome using the top-associated eQTL for the exposure and the same single nucleotide polymorphism (SNP) in the outcome GWAS. Standard errors 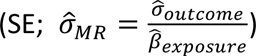 and 95% confidence intervals (CI) for *̂β_MR_* were calculated using the Delta method (56). Since eQTL effects were expressed as changes in gene expression (in standard deviation [SD] units) per additional effect allele, and the outcome effects were expressed on the log odds ratio (OR) scale, *̂β_MR_* can be interpreted as the change in the log(OR) of the outcome resulting from a 1-SD increase in gene expression. ORs were obtained by exponentiating the effect estimates.

For the primary analysis of AN and BE, we applied a Bonferroni-corrected significance threshold of p < 0.005 (0.05/(5×2)), accounting for the five validated *cis*-eQTLs and two outcome traits. As topiramate has also been reported to affect psychiatric disorders, we performed exploratory analyses of eight psychiatric traits: ADHD, ANX, AUD, BD, MDD, PD, PTSD, and SCZ. For these analyses, we applied a Bonferroni-corrected significance threshold of p < 0.00125 (0.05/(5×8)). Associations with p < 0.05 were considered suggestive. For all SMR results, we reported ORs, 95% CIs, and p-values.

### Sensitivity analysis

To assess the robustness of the SMR results and distinguish associations consistent with a shared causal variant from those potentially attributable to linkage disequilibrium (LD), we conducted three sensitivity analyses: SMR-multi (57), heterogeneity in dependent instruments (HEIDI) test (54), and Bayesian colocalization analyses (58).

Unlike SMR, which relies on single-variant estimates, SMR-multi is an extension of the SMR method that incorporates multiple *cis-*eQTLs rather than relying solely on the top-associated *cis*-eQTL (57). This method selects all SNPs with p < 5×10^-8^ within the *cis* region (i.e., within 500kb of the probe) and then removes SNPs in high LD with the top-associated SNP (i.e., LD r^2^ > 0.9). The causal effect was estimated for each SNP and then combined into a single test using an approximate set-based test (59) to account for LD between the SNPs. Only SNPs available in both the exposure and outcome datasets could be included; therefore, the number of instruments used in SMR-multi could vary across gene expression–outcome pairs.

The HEIDI test assesses whether the observed association between gene expression and the outcome is consistent with a shared causal variant rather than two distinct causal variants in LD. A significant HEIDI test (p_HEIDI_ < 0.05) indicates evidence of heterogeneity and is inconsistent with a single shared causal variant. We performed HEIDI using the following parameters as specified in Zhu et al. (54): p_eQTL_ < 1.57×10^-13^ (equivalent to a chi-squared value χ^2^_1_ = 10), LD pruning thresholds for removing SNPs in very strong LD (r^2^ > 0.9) or in very low LD (r^2^ < 0.05). LD was estimated from European-ancestry individual-level SNP genotype data from the 1000 Genomes Project (60).

Colocalization analyses were performed to assess whether gene expression and outcome associations within a locus were consistent with a shared causal variant. Five mutually exclusive hypotheses were evaluated: H0) no association with the exposure or the phenotype; H1) association with the exposure and not the outcome; H2) association with the outcome and not the exposure; H3) association with the outcome and the exposure by independent SNPs; H4) association with the outcome and the exposure by one shared SNP. The posterior probabilities (PP) of H0–H4 sum to 1, with PP of H4 (PP_H4_) > 0.70 considered evidence supporting colocalization (61). We performed colocalization analyses using coloc.abf() in the R package coloc (version 5.2.3) (58).

SMR, SMR-multi, and HEIDI analyses were performed using the SMR software tool (v1.4.2) (54). All other statistical analyses were performed in R (version 4.4.2), and all tests were two-tailed. These analyses were performed with the University of Queensland Research Computing Centre’s Bunya supercomputer (62).

### Power calculation

Statistical power was calculated to determine the minimum detectable effect size for each validated topiramate target across all psychiatric outcomes. We used the mRNd tool (63) (https://shiny.cnsgenomics.com/mRnd/) to estimate the minimum detectable effect size at 80% power for each gene expression-outcome pair. Type I error rates (α) were set at 0.05, 0.005 and 0.00125, corresponding to the nominal, primary, and exploratory significance thresholds used in this study.

## Results

We identified the genetic instruments (p_eQTL_ < 5×10^-8^) for 13 genes from brain *cis*-eQTLs and 15 genes from blood *cis*-eQTLs. To identify valid genetic instruments, we aligned the effect alleles to ensure that the direction of expression of all genes mimicked the agonistic or antagonistic action of topiramate and validated them by their association with a decreased risk of epilepsy (p < 0.05). This resulted in 2 validated genetic instruments from brain *cis*-eQTLs (*CACNB3*: rs2453469; *CA4*: rs117154637) and 3 from blood *cis*-eQTLs (*GRIK3*: rs11263973; *CA4*: rs34820870; GRIK1: rs2832236) to proxy for the pharmacological action of topiramate (**Table 2**). Those associated with an increased risk or showing no association were removed from further analyses.

**Table 2.**
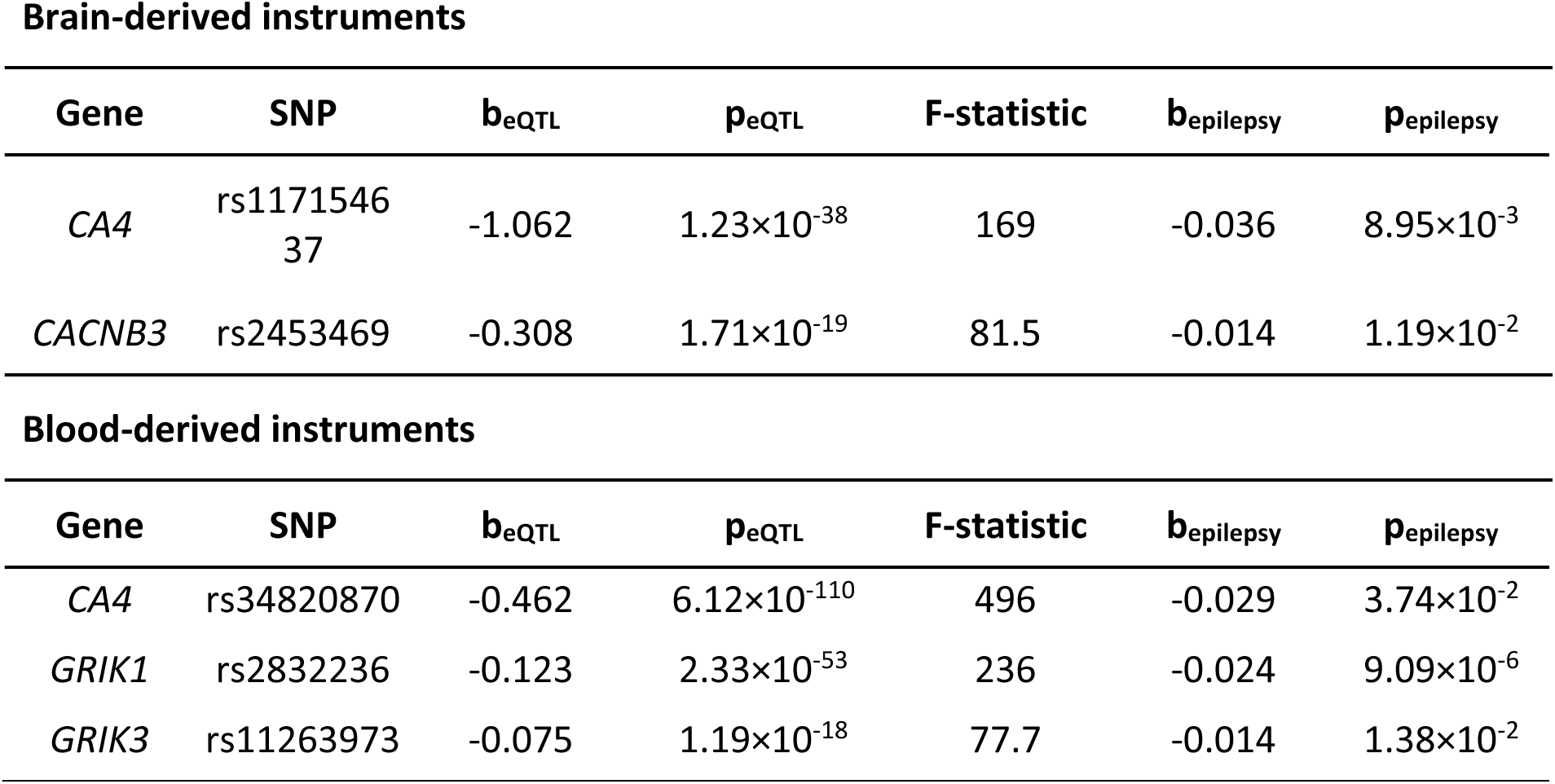
The validated genetic instruments to proxy for topiramate exposure in the brain or blood.

| **Brain-derived instruments** |  |  |  |  |  |  |
| **Gene** | **SNP** | **b<sub>eQTL</sub>** | **p<sub>eQTL</sub>** | **F-statistic** | **b<sub>epilepsy</sub>** | **p<sub>epilepsy</sub>** |
|---|---|---|---|---|---|---|
| *CA4* | rs117154637 | -1.062 | 1.23×10<sup>-38</sup> | 169 | -0.036 | 8.95×10<sup>-3</sup> |
| *CACNB3* | rs2453469 | -0.308 | 1.71×10<sup>-19</sup> | 81.5 | -0.014 | 1.19×10<sup>-2</sup> |
| **Blood-derived instruments** |  |  |  |  |  |  |
| **Gene** | **SNP** | **b<sub>eQTL</sub>** | **p<sub>eQTL</sub>** | **F-statistic** | **b<sub>epilepsy</sub>** | **p<sub>epilepsy</sub>** |
| *CA4* | rs34820870 | -0.462 | 6.12×10<sup>-110</sup> | 496 | -0.029 | 3.74×10<sup>-2</sup> |
| *GRIK1* | rs2832236 | -0.123 | 2.33×10<sup>-53</sup> | 236 | -0.024 | 9.09×10<sup>-6</sup> |
| *GRIK3* | rs11263973 | -0.075 | 1.19×10<sup>-18</sup> | 77.7 | -0.014 | 1.38×10<sup>-2</sup> |

### SMR analyses of AN and BE

After multiple-testing correction (p < 0.005), genetically predicted inhibition of blood *CA4* expression was significantly associated with a reduced risk of BE (OR = 0.867; 95%CI = 0.792 – 0.950; p = 0.002). This association remained significant in the SMR-multi analysis (p_SMR-multi_ = 4.9×10^-3^), which included three IVs (**Supplementary tables 2 & 3**). This association showed no evidence of heterogeneity in the HEIDI test (p_HEIDI_ = 0.536). Colocalization provided moderate support for a shared causal variant underlying *CA4* expression and BE (PP_H4_ = 0.58; **Figure 3A**), although this was below the prespecified threshold of PP_H4_ = 0.70. Conversely, we found no significant associations between any topiramate proxies and AN, even at the relaxed nominal significance threshold (P>0.05) (**Figure 2**).

**Figure 2.**
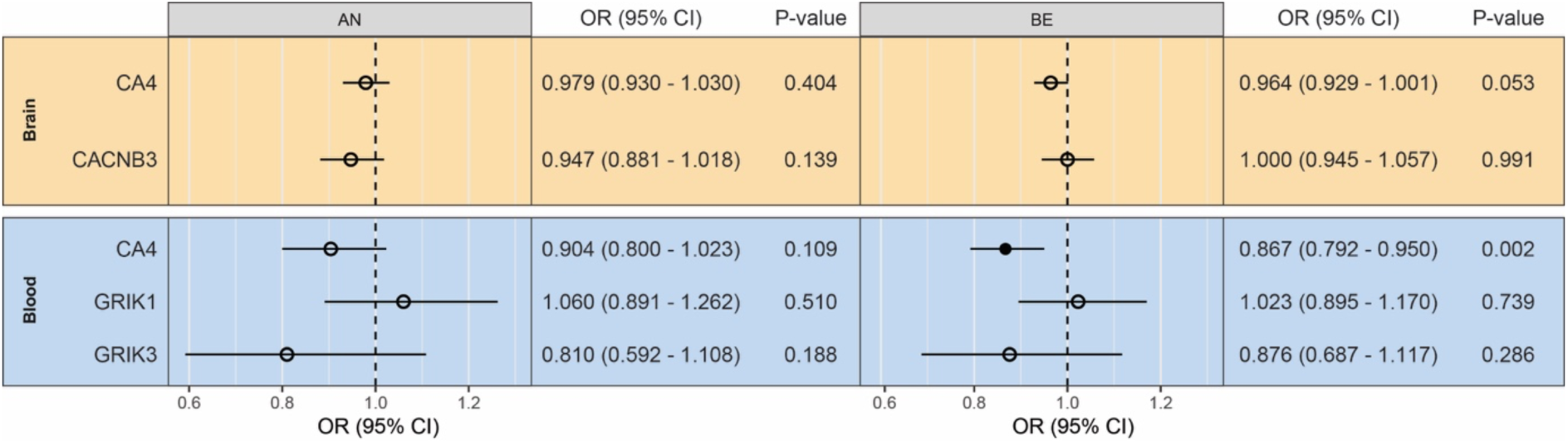
SMR analysis of genetically predicted expression of topiramate molecular targets for AN and BE risk. Brain-derived and blood-derived instruments are labelled in orange and blue, respectively. Odds ratios and 95% confidence intervals (CI) represent the changes in AN/BE risk per standard deviation decrease in gene expression. Estimates meeting the prespecified multiple-testing threshold (*P* < 0.005) are shown as filled points.

**Figure 3.**
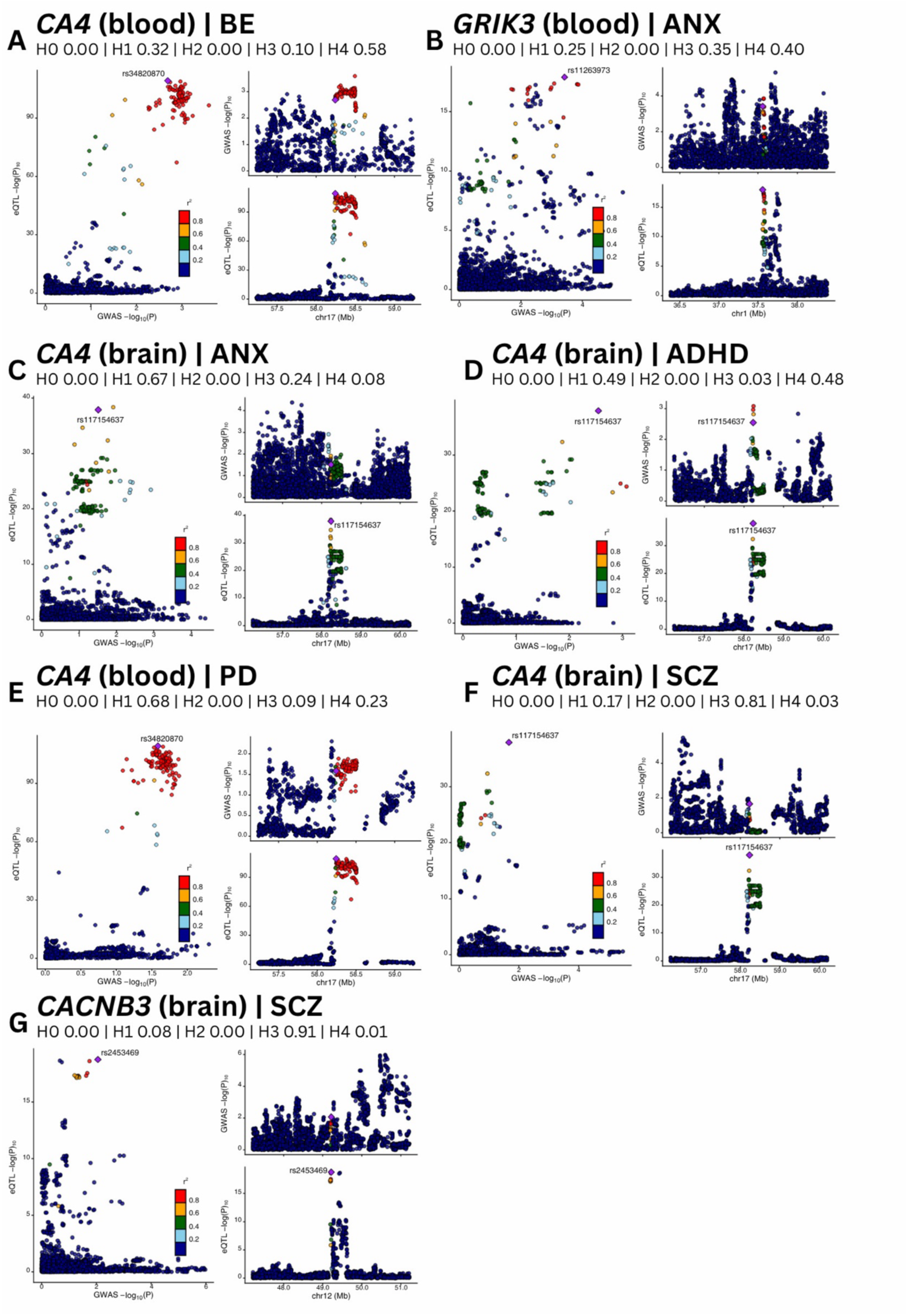
Regional genomic plots from colocalization analyses. (A) Colocalization between *CA4* expression in blood and BE. (B) Colocalization between *GRIK3* expression in the blood and ANX. (C). Colocalization between *CA4* expression in the brain and ANX. (D). Colocalization between *CA4* expression in the brain and ADHD. (E). Colocalization between *CA4* expression in blood and PD. (F). Colocalization between *CA4* expression in the brain and SCZ. (G). Colocalization between *CACNB3* expression in the brain and SCZ. The posterior probabilities of the five hypotheses (H0-4) for each analysis are shown.

### Exploratory SMR analyses

In the exploratory analyses with psychiatric traits as outcomes, after multiple-testing correction (p < 0.00125), we found a significant association between the genetically predicted inhibition of blood *GRIK3* expression and decreased ANX risk (OR = 0.752; 95%CI = 0.635 – 0.891; P = 0.001; **Figure 4**). The association remained significant in the SMR-multi sensitivity analysis, which included an additional IV (p_SMR-multi_ = 1.03×10^-4^). However, the association showed significant heterogeneity in the HEIDI test (p_HEIDI_ = 0.002), which indicates that the association is likely driven by distinct causal variants in LD rather than acting through a shared variant. At the same time, colocalization provided inconclusive evidence for a shared causal variant (PP_H3_ = 0.35; PP_H4_ = 0.40; **Figure 3B**). Therefore, these findings do not provide consistent support for a shared causal variant underlying *GRIK3* expression and ANX.

**Figure 4.**
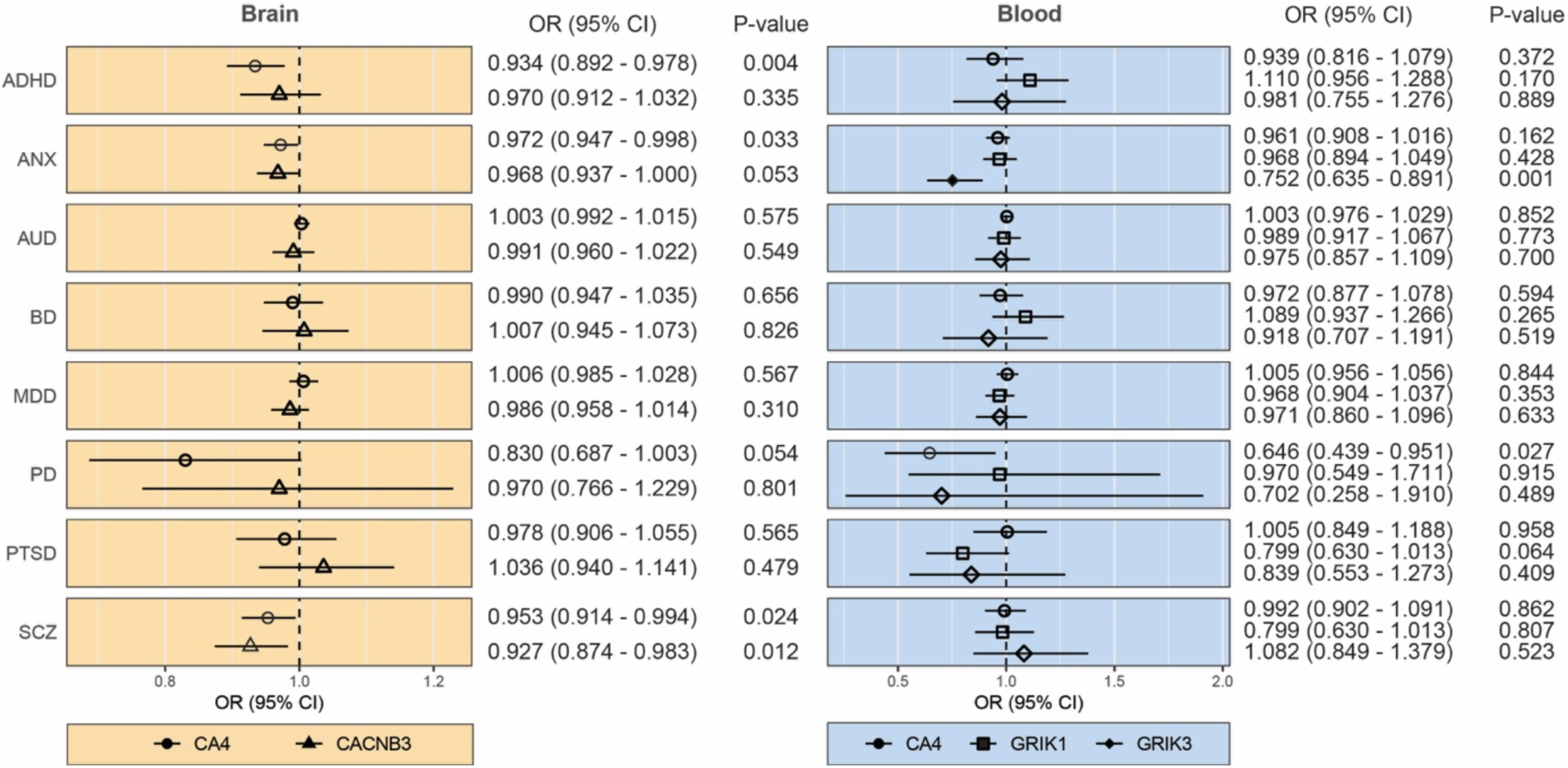
SMR analysis of the genetically predicted expression of topiramate molecular targets and eight psychiatric disorders. SMR results using brain (left) and blood (right) gene expression as the exposure. Odds ratios and 95% confidence intervals (CI) represent the changes in psychiatric disorder risk per standard deviation decrease in gene expression. Estimates with *P* < 0.00125 have filled points.

We found no further associations between genetically predicted topiramate actions and the risk of the eight psychiatric disorders after multiple-testing correction (**Figure 4**). Nominal associations were observed for ADHD, ANX, PD and SCZ (p < 0.05, **Supplementary Table 4**). These included associations of brain *CA4* expression with ADHD and ANX, blood CA4 expression with PD, and brain CA4 and CACNB3 expression with SCZ. However, support from sensitivity analyses was inconsistent: some associations attenuated in SMR-multi, showed HEIDI heterogeneity (**Supplementary Table 4**) or lacked support from a shared causal variant in colocalization analyses (**Figure 3C-G**).

### Power calculations

**Supplementary Table 5** summarizes the minimum detectable causal effect sizes at 80% power. Statistical power varied across gene expression-outcome pairs, reflecting differences in both instrument strength and outcome GWAS sample size. *CA4* expression in the brain had the greatest statistical power as its instrument explained the largest proportion of variance in gene expression (R^2^ = 0.097), followed by *CACNB3* in the brain (R^2^ = 0.046), *CA4* in blood (R^2^ = 0.016), *GRIK1* in blood (R^2^ = 0.008), and *GRIK3* in blood (R^2^ = 0.002).

For the primary outcome, AN, statistical power was greatest for the two brain-derived instruments. At the prespecified significance threshold for the primary analysis (α = 0.005), the minimum detectable ORs at 80% power were 1.076 and 1.111 for *CA4* and *CACNB3* expression in the brain, respectively. Power was more limited for the weaker blood-derived instruments, with minimum detectable ORs of 1.191, 1.274, and 1.480 for *CA4*, *GRIK1*, and *GRIK3*, respectively (**Supplementary Table 5**). Power was slightly greater for BE, with minimum detectable ORs of 1.060 and 1.088 for *CA4* and *CACNB3* expression in the brain, respectively (α = 0.005). Although AN and BE had similar total sample sizes, the BE GWAS had a higher proportion of cases, which contributed to greater statistical power.

For the exploratory outcomes (α = 0.00125), power similarly varied according to instrument strength and outcome GWAS sample size. For example, using the weakest instrument, *GRIK3* expression in blood, the minimum detectable OR was 3.263 for PD, compared with 1.273 for AUD and 1.264 for MDD. In contrast, the stronger brain-derived instruments provided substantially greater power; for AUD, the minimum detectable ORs were 1.042 and 1.062 for *CA4* and *CACNB3* expression in the brain, respectively (**Supplementary Table 5**).

## Discussion

This study applied drug-target MR to evaluate the potential causal effects of topiramate on AN and BE risk. Using validated genetic proxies for topiramate’s pharmacological targets, we found evidence that genetically predicted inhibition of *CA4* expression in blood was associated with a lower risk of BE, although colocalization provided only moderate support for a shared causal variant underlying this relationship. Nevertheless, these results warrant further follow-up on the effectiveness of topiramate in reducing BE and the potential role of CA4 in this relationship. In contrast, no robust evidence was found to suggest that the genetically predicted modulation of these targets influenced the risk of AN or the eight exploratory psychiatric outcomes.

CA inhibition is one of the proposed antiepileptic mechanisms of topiramate. CAs are ubiquitously expressed metalloenzymes that catalyse the reversible hydration of carbon dioxide to bicarbonate, contributing to pH homeostasis and small molecule transport (64–66). The 16 CA isoforms vary in their cellular location (cytosolic, membrane-bound, mitochondrial or secreted), and the inhibition of some isoforms has been suggested to suppress epileptic activity, although the exact mechanism remains poorly understood (65). One hypothesis is that the inhibition of neuronal CAs decreases intracellular bicarbonate concentrations, consequently reducing GABAergic depolarization associated with seizure susceptibility (67). Consistent with this hypothesis, topiramate has been shown to lower the steady-state pH of CA3 hippocampal neurons, which may contribute to its anticonvulsant effects (68–70). Although topiramate has been found to be a more potent inhibitor of CA2 and CA4 compared to other isoforms (71), inhibition of these isoforms does not appear to be strongly correlated with anticonvulsant activity (72).

CA4 was the only isoform for which we identified a genetic instrument suitable for SMR analysis. Compared with other CAs, the endogenous role of CA4 remains poorly characterized, with no endogenous role as a transporter or receptor reported to date (64). In this study, the genetically predicted inhibition of *CA4* expression in blood was robustly associated with reduced BE risk, surviving multiple-testing correction (p < 0.005) and remained significant in SMR-multi. However, colocalization analysis provided only moderate support for a shared causal variant underlying *CA4* expression and BE (PP_H4_ = 0.58). Furthermore, the association with *CA4* expression in the brain did not reach nominal significance (p > 0.05), although the 95% CIs overlapped with those for blood *CA4*. This apparent tissue-specific difference should therefore be interpreted cautiously and requires further investigation.

The role of CAs in BE remains largely unexplored. However, zonisamide, another anticonvulsant drug with CA inhibition properties, has also been suggested as a potential BE treatment, supporting the potential role of CAs in BE risk, although concerns over neurological and gastric side effects have been reported (73). CA4 has also been implicated in chemosensation (74–76). CA inhibition with acetazolamide has been reported to alter taste perception, particularly the perception of carbonation, in a case study (75). Meanwhile, another study demonstrated that pharmacological inhibition of extracellular CA4 substantially reduced carbon dioxide taste responses mediated by sour-sensing taste cells, with CA4 identified as the principal carbon dioxide taste sensor (77). Together, these findings suggest that CA inhibition may influence eating behaviour through changes in food perception and, consequently, food intake. However, the specific role of CA4 in BE and the mechanisms underlying this association require further investigation.

The absence of robust evidence for an effect on AN does not exclude an effect of topiramate on appetite or body weight. Weight loss is a relatively common pharmacological effect of topiramate, whereas AN is a complex psychiatric disorder characterized by behavioural and psychological features beyond reduced appetite and weight loss. Clinical reports suggest that topiramate-associated appetite suppression and weight loss may trigger or exacerbate AN-related symptoms in susceptible individuals (18–20), consistent with the possibility that the mechanisms driving weight loss may be distinct from those underlying susceptibility to AN. Recent genetic evidence further indicates that AN and BE, despite substantial shared genetic architecture, differ in their relationships with anthropometric, metabolic, and impulse-control traits (26). In particular, BE shows stronger genetic relationships with impulse-control-related traits (26,78), whereas AN has a distinct metabolic component supported by genetic and metabolomic evidence (26,79,80). Thus, genetic liability to AN may not fully capture the mechanisms underlying topiramate-associated appetite suppression or weight reduction. Overall, these findings highlight the importance of studying eating disorders as distinct phenotypes to better disentangle the diverse biological and behavioural pathways through which drugs and other exposures may influence their risk.

In clinical populations, the relationship between topiramate use and weight-related outcomes may also be influenced by indication, comorbidities, and concomitant medications. A US national survey conducted from 2013 to 2016 found that patients treated for epilepsy used an average of 5.3 medications, with medication burden increasing with age and chronic comorbidities (81). Such complexity makes it difficult to disentangle the effects of topiramate from those of other medications and comorbid conditions in observational studies and further highlights the distinction between the target-specific effects investigated here and the effects observed during clinical treatment. Future studies incorporating information on treatment indication, comorbidities, and concomitant medications may help clarify the factors contributing to topiramate-associated weight loss.

Exploratory analyses provided no robust evidence that genetically predicted modulation of the validated topiramate targets influenced the eight additional psychiatric outcomes. These analyses were motivated by the high burden of comorbidity among individuals with epilepsy (82). Genetically predicted inhibition of blood *GRIK3* expression was associated with reduced ANX risk and survived correction for multiple testing; however, this association showed significant heterogeneity in the HEIDI test, while colocalization provided inconclusive evidence for a shared causal variant. Further nominal associations were observed for ADHD, PD, and SCZ, but none survived correction for multiple testing, and support from sensitivity analyses was inconsistent. Overall, these findings require cautious interpretation and independent validation. Notably, topiramate has also been investigated for preventing or reversing weight gain induced by psychotropic medications (83–85), further highlighting that its effects on body weight may vary according to clinical context.

Several limitations need to be considered. First, genetically predicted differences in gene expression represent proxies for lifelong variation in target expression and do not directly reproduce the magnitude, timing, or biochemical mechanism of pharmacological modulation by topiramate. Although our positive-control analysis selected instruments whose effects on epilepsy were directionally consistent with topiramate’s therapeutic action, this does not establish that altered gene expression is functionally equivalent to drug-induced activation or inhibition of the encoded protein. Furthermore, eQTLs may not fully capture variation in functional protein abundance, as mRNA and protein levels are not always strongly correlated (86,87), including in the human brain (88). Proteome-wide association data from brain tissue remain limited by sample size and technical constraints, although ongoing advances in proteomics and large-scale biobank initiatives may enable more comprehensive protein-based analyses in the future (89,90). Second, our analyses were restricted to populations of European ancestry, limiting the generalizability of our findings to other ancestral populations. Future studies using more genetically diverse datasets are needed to determine whether these findings extend to other populations. Third, our analyses were restricted to topiramate’s established protein targets for which suitable genetic instruments could be identified and validated against epilepsy risk. The relative contributions of these targets to topiramate’s pharmacological effects remain incompletely understood, and some effects on neurotransmitter signalling may arise indirectly through modulation of other molecular pathways. Consequently, effects mediated through targets for which suitable instruments were unavailable, or through downstream pathways not captured by our approach, could not be evaluated. Given that topiramate interacts with multiple molecular targets, its effect on AN, BE and other psychiatric outcomes may involve other target-mediated or off-target mechanisms that are difficult to investigate using our approach, particularly when suitable positive-control outcomes for validating genetic proxies are lacking. Additionally, genetic instruments validated for topiramate’s therapeutic effects on epilepsy may not fully capture its effects in other clinical indications, including migraine (91). Further research is needed to clarify its effects across different target pathways and patient populations.

This study has several strengths. The application of drug-target MR enabled us to investigate a potential adverse effect of topiramate that is difficult to evaluate using conventional study designs. The use of genetic instruments derived from large GWAS and eQTL datasets help mitigate some of the challenges inherent to observational and human drug intervention studies, including confounding, limited sample sizes, and ethical constraints. Incorporating eQTLs from both brain and blood also allowed us to investigate genetically predicted target perturbation across central and peripheral tissues. Power analyses further helped contextualize the null findings. The stronger brain-derived instruments, particularly those for *CA4* and *CACNB3*, provided good power to detect smaller effects on AN and several larger exploratory GWAS. However, power was substantially lower for weaker blood-derived instruments, meaning that clinically relevant effects through these targets cannot be excluded. Although MR does not replace experimental validation in mammalian studies, our findings provide genetic evidence that can be triangulated with evidence from other study designs to better understand the potential effects of topiramate on AN and BE risk. Functional studies in model organisms, including ongoing work using *Caenorhabditis elegans* to characterize the anorexic (appetite-suppressing) effects of topiramate (92), may provide complementary evidence for identifying biological pathways underlying topiramate-associated changes in appetite and body weight.

Overall, this study found suggestive evidence that genetically predicted inhibition of *CA4* expression in blood was associated with reduced BE risk. In contrast, no robust evidence that genetically predicted modulation of validated topiramate targets influences AN risk was identified. Exploratory analyses similarly provided no robust evidence for effects on ADHD, ANX, AUD, BD, MDD, PD, PTSD, or SCZ. These findings warrant further triangulation with clinical, pharmacological, and functional evidence to clarify the mechanisms underlying topiramate-associated appetite suppression and weight loss and their relevance to eating-disorder risk.

## Data Availability

All data produced in the present work are contained in the manuscript.

## Data availability

Publicly available eQTL datasets and GWAS summary statistics were used in this study. GWAS summary statistics are available from their respective publications or the Psychiatric Genetics Consortium: https://pgc.unc.edu/for-researchers/download-results/. BrainMeta eQTL summary statistics are available at https://yanglab.westlake.edu.cn/software/smr/#DataResource. Blood-derived *cis*-eQTL from eQTLGen phase I are available at: https://www.eqtlgen.org/cis-eqtls.html.

## Code availability

Bash and R scripts used for analysis are available from the corresponding author upon reasonable request.

## Funding

L.D.H. was supported by an Australian Research Council Discovery Early Career Researcher Award (DE240100014). E.Y.C. was supported by scholarships from the University of Queensland.

## Author contributions

K.N. and E.Y.C. contributed equally to the project. K.N., E.Y.C. and J.C.J. performed the analyses. K.N. and E.Y.C. drafted the manuscript. J.C.J. supervised the project. L.D.H. conceived and designed the study and provided overall supervision. All authors interpreted the data, reviewed and approved the final manuscript.

## Ethics declarations

### Competing interests

The authors declared no competing interests.

## Ethics declarations

This project received ethical approval from the Institutional Human Research Ethics Committee, University of Queensland (Project Number: 2024/HE001677).

## Notes

### Competing Interest Statement

The authors have declared no competing interest.

### Author Declarations

This study only used publicly available summary statistic data. All sources are provided in the manuscript.

